# Predicting Growth of COVID-19 Confirmed Cases in Each U.S. County with a Population of 50,000 or More, Revised July 8, 2020

**DOI:** 10.1101/2020.05.29.20117044

**Authors:** Leon S. Robertson

**Affiliations:** Yale University School of Public Health

## Abstract

A simple model of local spread of COVID-19 is needed to assist local governments and health care providers prepare for surges of clinical cases in their communities. National and state based models are inadequate because the virus is introduced and spreads at different rates in local areas. In the U.S. as of July 3, 2020, 73 percent of cases and 84 percent of deaths occurred in the 200 counties with the most cases and deaths. Each county has its own function of cases in time that can be used to predict increases in reported cases two weeks in advance for each of 988 counties in the U.S. with populations of 50,000 or more inhabitants. A logarithmic model based on growth in cases during the past 30 days is substantially predictive of increase in cases during the subsequent 14 days. Predicted increase in cases for the 988 U.S. counties will be published online daily.

## INTRODUCTION

Projected COVID-19 cases and deaths from various mathematical models using different methods initially produced quite different results in the U.S. because of the variance in assumptions about how the virus and people would behave. As more data became available the projections converged somewhat but still varied substantially [1]. The early projections did serve the purpose of motivating politicians, however belatedly, to adopt policies to slow the spread of the virus. In the U.S. the timing of stay-at-home (shutdown) orders varied among the states. The business operations and other gatherings prohibited during the shutdown also varied somewhat among the states but all left plenty of leeway for the virus to continue to spread. Several state governors did not issue such orders and many announced partial or complete termination of the orders in late April and early May, 2020. A few prohibited local governments from continuing the shutdown or requiring masks and physical distancing. In Wisconsin and Oregon the order was voided by judges. Analysis of numerous factors in relation to cases and deaths indicate that the shutdowns reduced deaths by about half and cases about 48 percent [2].

Some state and local governments issued standards for physical distancing in businesses and other organization as well as wearing masks in public places after the shutdown but enforcement will be problematic as it was in states with stay-at-home orders. If local medical care facilities are to be prepared for an influx of cases, it is important to be able to predict the rise in numbers in local areas as early as possible. Large swaths of counties in the U.S. have no hospital beds, and if they have them, no intensive care units. More than half of U.S. counties have no intensive care beds [3]. A COVId-19 patient can occupy an intensive care bed for weeks before dying or leaving the hospital alive.

Others have recently developed short-term models based on estimates of transmission rates by county but these models are very difficult to use by busy local authorities and health care professionals (4–6). Estimating the transmission rate of the COVID19 virus is problematic because of its behavior. Many people who are infected do not experience significant or any symptoms but shed virus that infects others in physical proximity or in contact with surfaces where it dwells for a time. For example, tests of a sample of the population of Westchester County New York, the first “hot spot” in that state, indicated that 16.7 percent of people had antibodies to COVID-19 [7]. If the sample is representative of the population, 161,673 people in Westchester County had been exposed to enough of the virus to produce antibodies (.167 x 967,506 people in the population) but there were only 31,294 “confirmed cases” reported by the County Health Department as of May 10, 2020. Some 81 percent of those who may be positive for antibodies but not reported as cases either experienced mild enough symptoms to avoid seeking help or no symptoms at all. In New York City, 21 percent of the sample had antibodies but in Bronx County only about 13.8 percent of those had turned up in the “confirmed cases” count by May 10, about 86 percent did not. The difference between Westchester and Bronx counties could be a result of fluctuations in sampling or they could represent differences in help-seeking among people with relatively high (Westchester) or low (Bronx) incomes or other differences among the populations. Whether people with antibodies are immune to future infections, and if so for how long, is unknown. These characteristics of virus spread alone make the application of traditional epidemic modeling difficult. These models require information on infection and recovery rates as well as immunity [8] which involves testing vast numbers of people.

Human behavior is obviously a major factor in the spread of a human transported infectious disease. Human responses to the news that the COVID-19 virus was spread by breathing, coughing, sneezing, talking and singing as well as touching surfaces by those infected varied from substantial risk avoidance behavior (e.g., reducing physical proximity to other people, frequent hand washing, wearing face masks) to mockery of those who did so and protests against requirements to do so [9]. Photographs and videos of street protesters against shutdowns showed many people in close proximity to one another with no face masks [10.

Travel data based on tracing cell phone movements indicate that travel decreased prior to the adoption of stay-at-home orders in many metropolitan areas but increased in time later [11] suggesting that risk avoidance began before the orders and deteriorated thereafter. Since manifestation of symptoms lags infection by the virus for some two weeks, data projecting numbers two weeks in advance could give local areas at least a warning of acceleration in cases assuming that the data were available promptly. It turns out that a model based on that idea is a better predictor of future cases than cell phone movement.

The original version of this paper showed that a model based on days since the first confirmed case in a given county was effective at predicting total accumulated cases 14 days in advance. The average R^2^ among U.S. counties was 0.93 with none less than 0.70. As states reopened after the shutdowns, COVID-19 began to spread again, severely in states that opened more abruptly. The model became less predictive, particularly in counties that phased in reopening based on benchmarks for virus control. The purpose of this paper is to report a revised model using the most recent thirty day trend in cases to predict new cases in each county 14 days in advance.

## METHODS

I used ordinary least squares regression of the trend in the logarithm of confirmed cases to predict the number of cases two weeks in the future. The study was limited to counties with 50,000 or more population to avoid random variation in small numbers. The hypothesized predictive equation for 30 days prior to a given date is log(accumulated cases_t+14_)=a +b[days_t-30 to t-1_). To smooth the daily variation in cases due to differential reporting on weekends and holidays, “cases” is the seven day moving average of cases for each county. The 14 days were added to account for the average time between exposure and manifestation of symptoms. Daily numbers of accumulated confirmed cases in each county through July 3, 2020 were downloaded from usafacts.org [12].

## RESULTS

Data on all variables were available for 988 counties. The total population of these counties (278,567,774) was about 85 percent of the estimated 2019 U.S. population. The number of cases in these counties was 2,439,951, about 88 percent of the U.S. total. The cases were concentrated than these numbers indicate. As of July 3, 2020, 73 percent of cases and 84 percent of deaths occurred in the 200 counties with the most cases and deaths.

The logarithmic model is effective at predicting total accumulated cases 14 days in advance. The average R^2^ was 0.93 with less than 5 percent below 0.70. Figure 1 shows the distribution of the coefficients. More than a third was less than one and 55 percent were less than two. Higher coefficients are indicative of a curve bending upward rapidly, far from the flattening needed to indicate containment of the virus. Figure 2 shows the patterns indicated by increased coefficients during a 60 day period. If 3 counties each started with 100 cases on a given day, but had the different curves shown in Figure 2, the accumulated cases after 60 days would be more than 6000 for the county with a coefficient of .07, more than 1000 for the county with a coefficient of .04 and 182 for the county with a coefficient of .01. Such is the nature of logarithmic growth. Although the .01 coefficient looks relatively benevolent, a coefficient must be zero for growth in cases to be flat.

**Figure 1.**
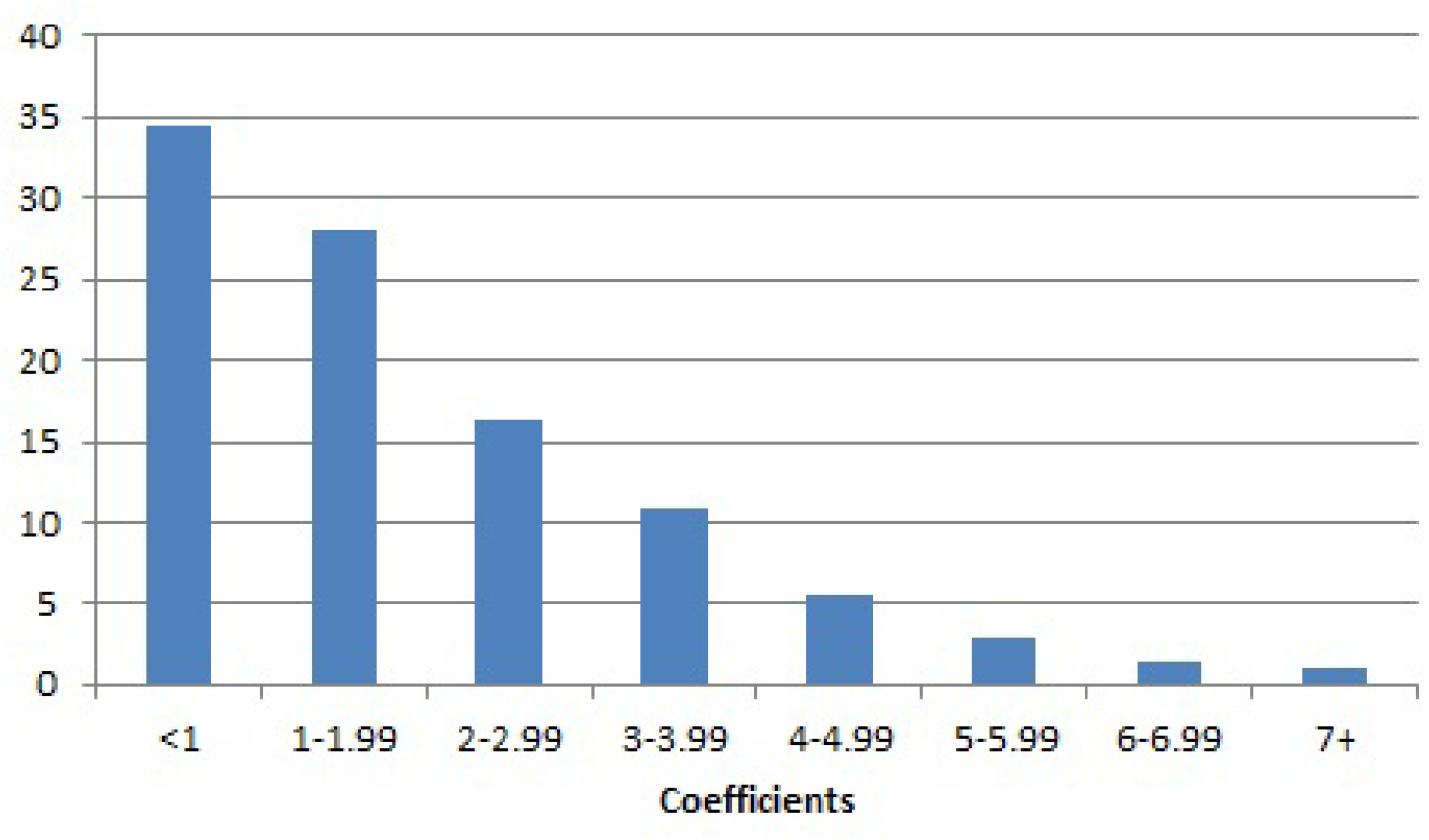
Percent of Regression Coefficients among U.S. Counties from the Logarithmic Model

**Figure 2.**
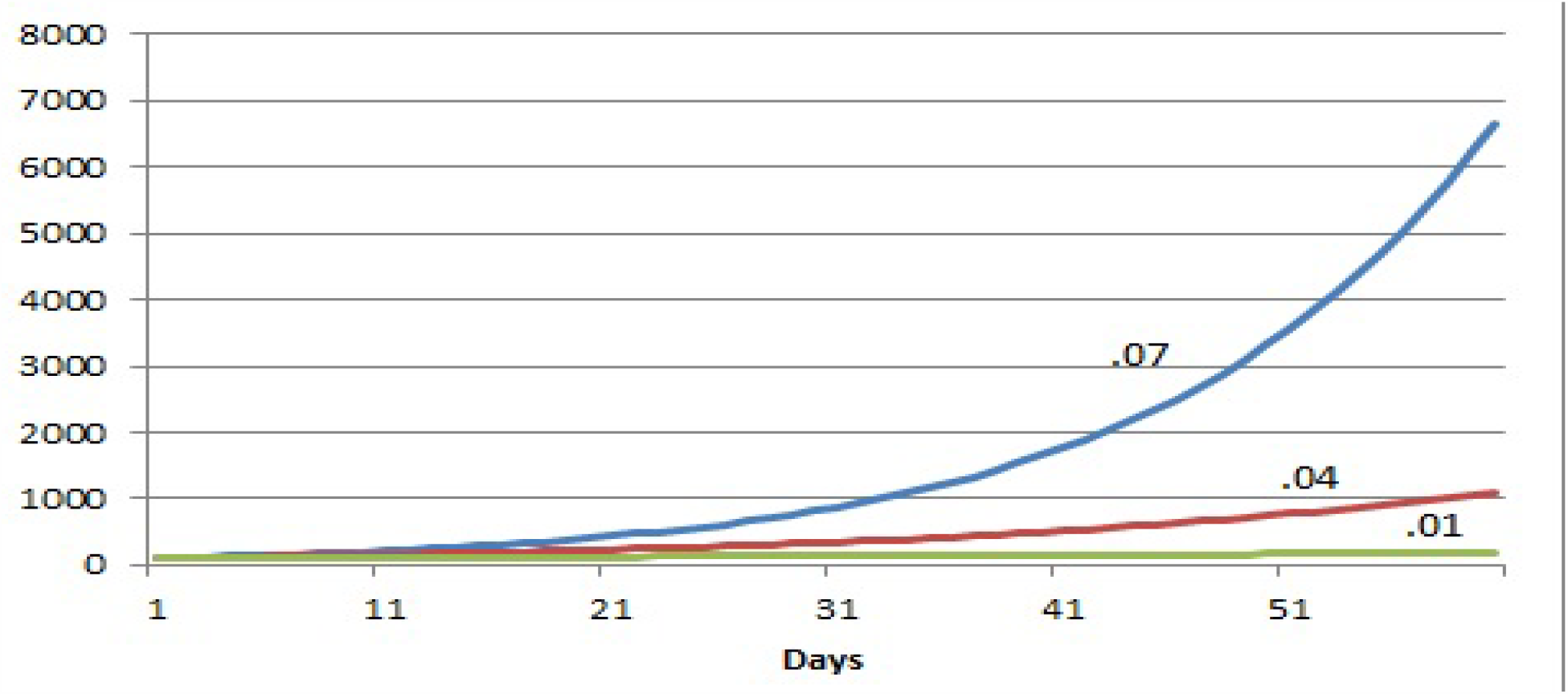
Slope of Accumulation of COVID-19 Cases When Regression Coefficients Are at Specified Levels

The fit of the model is illustrated in Figure 3. The number of actual accumulated cases July 3, 2020 closely fits the number predicted 14 days earlier.

**Figure 3.**
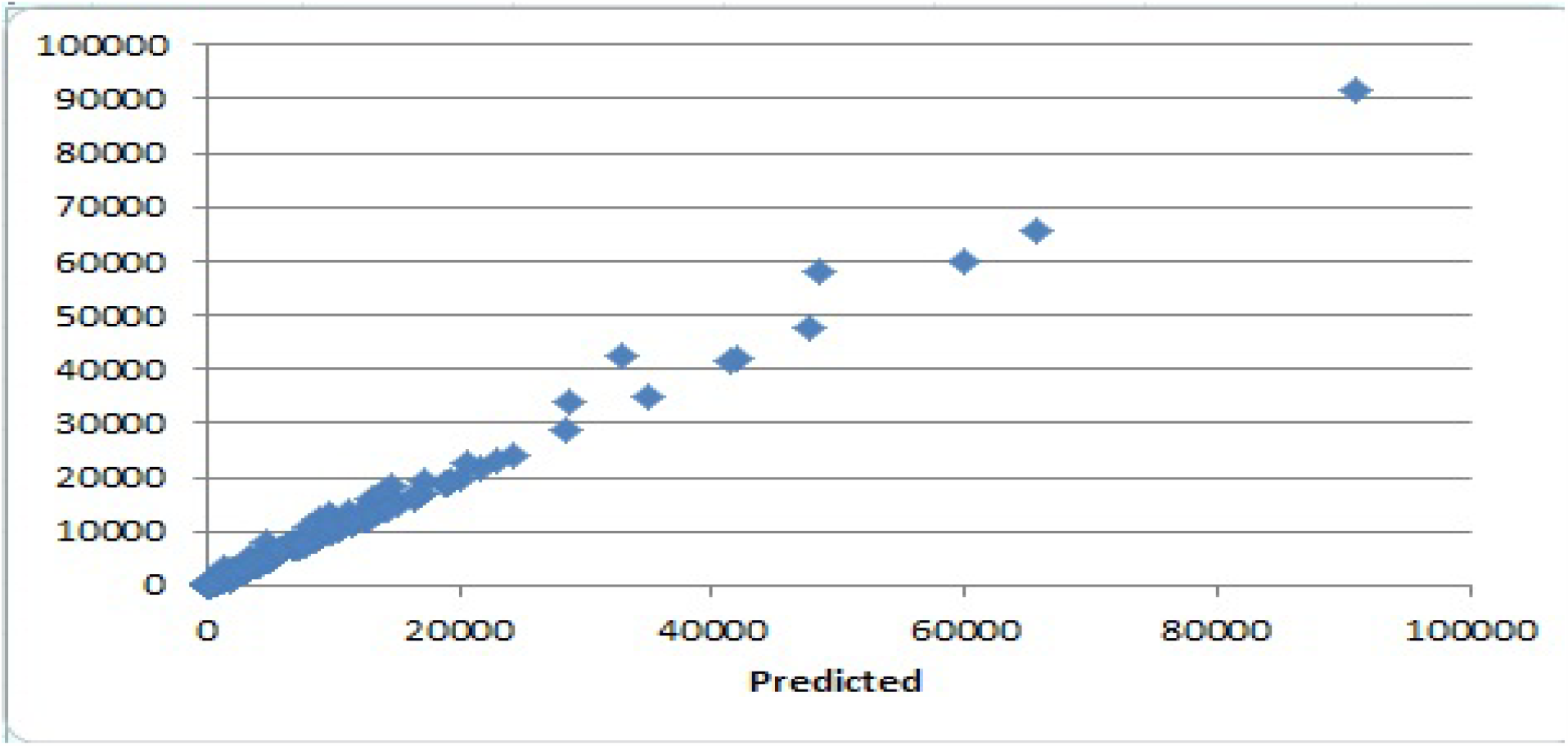
Actual Accumulated Cases in Each County and Number Predicted by July 3, 2020

## CONCLUSION

These data further illustrate that the spread of COVID-19 is very different in local areas. The data do not predict what will happen in the longer term but it can provide state and county officials and medical care facilities with estimated case counts 14 days in advance. The expected new cases two weeks in advance for each county will be updated daily. The results can be downloaded as a comma delimited file at www.nanlee.net. The data are listed by state and county in alphabetical order.

Without the shutdown, the COVID-19 virus would have caused more than enough severe illnesses to overwhelm the medical care system sooner in many cities in the U.S. As for rural America, low density reduces the individual risk but the medical system in many counties is without hospitals and those that have them are often without intensive care beds. As of this writing the virus continues to surge in larger cities and is invading the less dense areas of the country. There is no evidence of a seasonal decline as there has been with other manifestations of corona viruses [13].

We do not know the degree to which those who survived infection will have immunity and for how long. We do not know how many people will be gravely ill and survive or die before the curve is flattened by changes in behavior or tracing and quarantine. The development of a successful vaccine in time to curb the exponential slope in cases and deaths in many counties is unlikely.

## Data Availability

References to the public datasets are in the paper as well as a link to a file of parameters for users of the method.

https:www.nanlee.net

